# Delay distributions limit the identifiability of rapid variations in epidemics

**DOI:** 10.64898/2026.07.23.26358599

**Authors:** Jingjing Tang, Bryan Wilder, Roni Rosenfeld

## Abstract

Public-health surveillance systems rely on downstream indicators to infer latent infection incidence, but delays and observation noise provide only an indirect and temporally distorted view of the underlying epidemic process. Reconstructing upstream epidemic trajectories from these observations is therefore an ill-posed inverse problem, in which different reconstruction assumptions may produce different trajectories that remain consistent with the observed data. Here, we develop a general spectral framework that quantifies the statistical distinguishability of candidate upstream trajectories under delayed and noisy observations. We show that epidemiological delay distributions impose a frequency-dependent limit on the information that surveillance observations retain about upstream epidemic dynamics. This limitation propagates to epidemiological inference, making some quantities substantially more sensitive to reconstruction assumptions than others and rendering distinct event-impact profiles difficult to distinguish from downstream observations.

## Introduction

Inference about epidemic dynamics commonly relies on observations such as confirmed cases, hospitalizations, and deaths to infer latent infection incidence [22, 29]. These observations are generated through biological progression, healthcare seeking, testing, and reporting, and therefore provide an incomplete and temporally distorted view of the underlying epidemic process [14, 21, 11]. The resulting observation process determines which temporal information about the underlying epidemic remains available for statistical inference (Fig. 1A–F). Information about rapid temporal variation is preferentially attenuated during the observation process, making short-timescale epidemic dynamics progressively less distinguishable than slower variations (Fig. 1D–H). Consequently, under a given delay and observation process, surveillance observations possess a finite temporal resolution beyond which distinct upstream epidemic trajectories become statistically indistinguishable, independent of the particular reconstruction algorithm employed. This raises a question that precedes reconstruction itself: which differences between epidemic histories are identifiable from the observations, and which remain underdetermined by the observations themselves?

**Figure 1:**
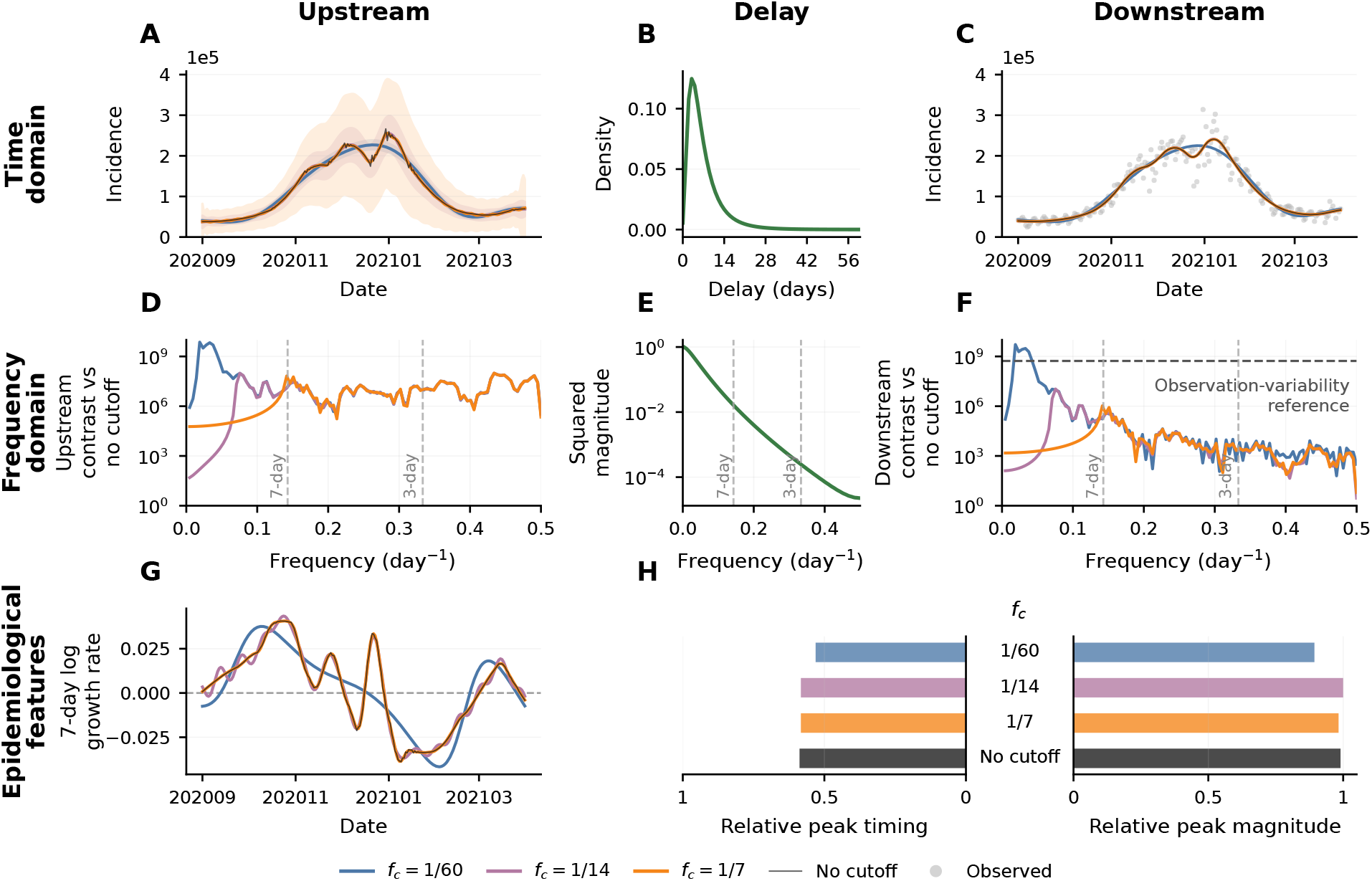
Delayed observations progressively reduce temporal information available for epidemic inference. Columns show the upstream incidence scale, the delay process, and the downstream surveillance scale. Each colored upstream reference is constrained to Fourier components with *f*≤ *f*_c_. (*A*) Frequency-constrained upstream reference trajectories estimated from U.S. confirmed-case incidence, with shaded bands showing locally non-identifiable perturbations. (*B*) COVID-19 symptom-onset-to-report delay distribution [33]. (*C*) Delay-convolved downstream reference trajectories and observed U.S. daily confirmed-case incidence [12]. (*D–F*) Upstream spectral contrast, squared delay-kernel magnitude, and downstream spectral contrast. (*G,H*) Seven-day log growth rates and relative peak summaries derived from the same reference trajectories. Colors denote cutoff-specific references; black denotes the no-cutoff reference, and gray points denote observed incidence. Displayed spectra omit the zero-frequency component. Construction details are provided in Appendices S2 and S3.

Existing work has primarily addressed this question by developing methods to reconstruct latent epidemic trajectories from delayed observations, including back-projection[4], deconvolution[18], Bayesian latent-incidence reconstruction[8], and renewal-based inference[10, 19]. Reconstructing latent incidence from delayed observations remains an ill-posed inverse problem because multiple upstream trajectories may be consistent with the same downstream observations[29]. Stable inference therefore requires regularization, prior distributions, or mechanistic constraints that select among multiple trajectories compatible with the observed data [29, 4, 25]. Posterior uncertainty quantifies what remains uncertain after introducing regularization and modeling assumptions, whereas observation-level identifiability asks what is determined by the observations themselves. This distinction motivates a broader question that extends beyond epidemic reconstruction itself.

Related concepts have also been developed in control theory and statistical detection theory. In control theory, observability characterizes whether latent states are recoverable from observations under specified dynamical and observation models[24, 27]. Statistical detection theory instead quantifies the distinguishability of competing signals or hypotheses in noisy observations using likelihood-based measures that underpin ideal-observer analyses in communication systems, signal processing, and medical imaging[20, 2, 1]. Neither framework directly characterizes the observation-level identifiability of epidemic trajectories under realistic delay distributions and observation noise. Addressing this gap requires linking epidemic delay processes and observation noise to likelihood-based measures of statistical distinguishability.

The epidemic observation process admits a natural spectral representation because convolution with a delay distribution becomes multiplication in the frequency domain[30, 6]. This representation directly characterizes how temporal information is preserved across frequencies, with realistic epidemic delay distributions preferentially suppressing high-frequency components before they appear in downstream observations (Fig. 1D–F)[8, 28]. Because statistical distinguishability depends on the information preserved by the observation process, observation-level identifiability is likewise expected to vary systematically across temporal scales.

Motivated by this observation, we develop a general framework for characterizing observation-level identifiability based on a spectral representation of delayed epidemic observations. By combining the frequency response of epidemiologically realistic delay distributions with observation-model-specific likelihood weights, we derive a separability functional that determines whether differences between upstream trajectories remain recoverable in downstream surveillance data. This formulation yields explicit identifiability limits across common observation models and characterizes which temporal features are recoverable from the observations themselves, independent of the particular reconstruction method employed.

## Methods

### Spectral representation for the delay model

Let *U* (*t*) denote the upstream process, such as latent infection incidence, and let *D*(*t*) denote a downstream surveillance observable, such as reported cases, hospital admissions, or deaths. The delay from upstream infection to downstream observation is described by a nonnegative delay density *g*(*τ* ), supported on *τ* ≥ 0 and normalized so that 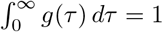. A conversion rate *p* ∈ (0, 1] represents the probability that an upstream event generates a downstream observation. One common example is the ascertainment rate, which describes the proportion of latent infections ultimately recorded as confirmed cases. Because testing- or reporting-related changes typically occur on coarse temporal scales [9, 13, 32] relative to the support of *g*, we treat *p* as locally constant.

We combine delay and conversion into the observation kernel

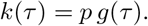

The downstream conditional mean is

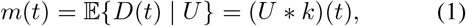

where ∗ denotes convolution,

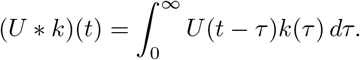

Equivalently, the downstream observation can be written as

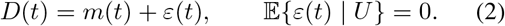

The observation model is illustrated in Fig. 1A–C.

For additive Gaussian observation models, *ε*(*t*) corresponds directly to Gaussian noise. For count-based observation models such as Poisson or negative-binomial sampling, equation (2) should instead be interpreted as a mean-plus-residual decomposition, where *ε*(*t*) denotes deviations from the conditional mean.

Because convolution in the time domain corresponds to multiplication in the frequency domain, it is natural to examine the conditional mean spectrally. Taking Fourier transforms gives

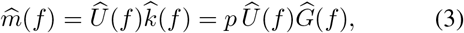

where 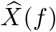 denotes the Fourier transform of *X*(*t*). When *p* = 1, equation (3) reduces to the delay-only setting without conversion loss. This frequency-domain representation highlights the attenuating effect of delay processes: for many epidemiological delay distributions, the magnitude |*Ĝ* (*f* )| decreases with frequency, thereby attenuating high-frequency upstream variation before it reaches the downstream mean. This attenuation mechanism is illustrated in Fig. 1D–F.

This framework extends naturally to multilayer delay structures in which several delay mechanisms act sequentially, such as incubation, testing, reporting, and hospitalization delays. Let *g*_1_, *g*_2_, …, *g*_L_ denote normalized delay kernels associated with successive stages, and let *p*_1_, *p*_2_, …, *p*_L_ denote the corresponding conversion rates. The overall conversion rate is 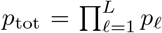, while the overall delay kernel is

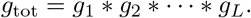

The overall observation kernel is

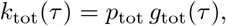

and the downstream conditional mean satisfies

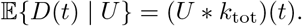

In the frequency domain, the corresponding transfer function factorizes as

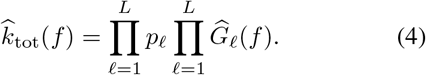

Thus, sequential delay mechanisms compound their spectral attenuation multiplicatively, progressively suppressing high-frequency upstream variation before it reaches the downstream process.

### Spectral separability of upstream trajectories

We quantify upstream identifiability by asking whether two candidate upstream trajectories *U*_0_ and *U*_1_ could be distinguished from the downstream observations. The statistical scale for this comparison is set by the downstream likelihood, but the effect of delay is most transparent after expressing the resulting likelihood separability in spectral coordinates. Let

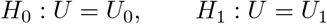

denote the two simple hypotheses, and let

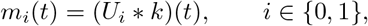

be the corresponding downstream mean trajectories. Their observable contrast is

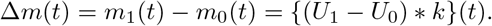

On a finite observation window, let **F** be the unitary discrete Fourier transform and let *K* be the diagonal transfer operator with entries 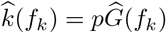. In the corresponding finite-window spectral representation,

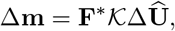

where ΔÛ is the Fourier transform of *U*_1_ − *U*_0_. Let *M* index the downstream observation model, and let **W**_*M*_ denote the likelihood weight induced by that observation model for the finite contrast between *m*_0_ and *m*_1_. This weight plays the role of observation precision: downstream contrasts in regions with greater observation variability receive smaller weight, and locally the weight reduces to inverse conditional variance. We define the spectral separability between *U*_0_ and *U*_1_ as the likelihood-weighted downstream contrast expressed in the upstream frequency domain,

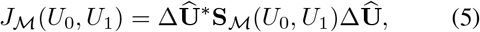

with

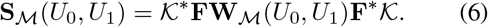

The transfer operator *K* attenuates upstream variation according to the delay distribution and conversion probability. After transforming back to the observation window, **W**_*M*_ scores the resulting downstream mean contrast on the scale of observation variability specified by the observation model. Consequently, *J*_*M*_ is small when upstream differences are concentrated at temporal scales suppressed by the delay kernel, or when the delay-filtered downstream differences are small on the observation-variability scale.

The same quantity can be written on the downstream mean scale as

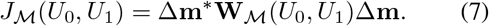

Thus *J*_*M*_ measures the part of the upstream spectral contrast that remains after delay filtering and is informative under the downstream observation model. **S**_*M*_ is a spectral operator that combines delay attenuation with the likelihood weight induced by the observation model. This form retains the intuition that identifiable upstream contrast must both survive delay filtering and remain large relative to observation variability. In special cases such as stationary Gaussian noise, the operator reduces to a frequency-by-frequency weight.

### Observation models and non-identifiability bound

We evaluated three observation models commonly used for epidemic surveillance data: additive Gaussian noise, Poisson counts, and negative-binomial counts. The corresponding likelihood weights are summarized in Table 1; full derivations are given in Appendix S1. For Gaussian observations with common covariance 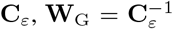. If the Gaussian noise is stationary, the Fourier basis diagonalizes this covariance and equation (5) becomes the familiar frequency-wise weighted contrast. For conditionally independent Poisson and negative-binomial counts, **W**_*M*_ is diagonal in time, with entries determined by the exact likelihood curvature between *m*_0_(*t*) and *m*_1_(*t*). The local approximations in Table 1 give the corresponding inverse-variance weights in the small-contrast limit.

**Table 1:**
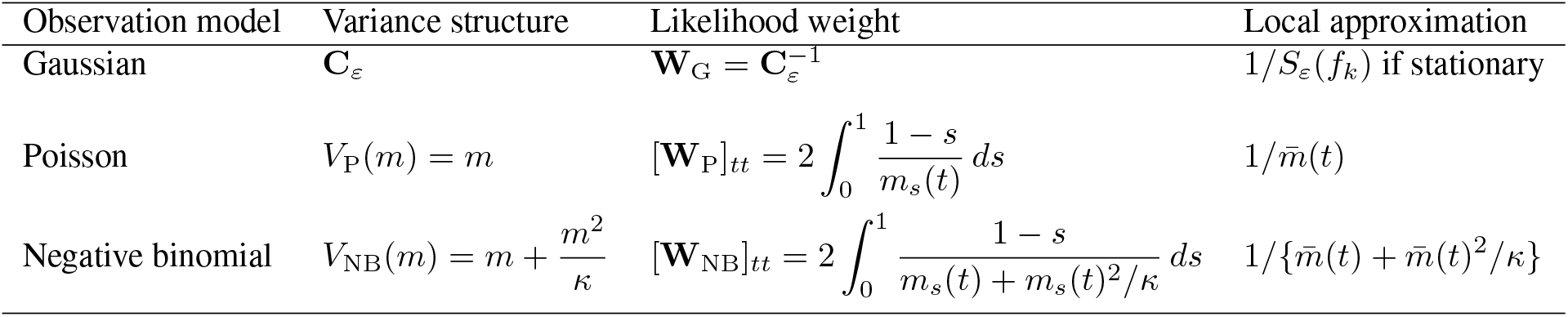
Likelihood weights for downstream separability. The separability can be written as *J*_*M*_ = Δ**m**^∗^**W**_*M*_Δ**m**, where Δ**m** is the delay-filtered downstream contrast. For count models, *m*_s_(*t*) = *m*_1_(*t*) + *s{m*_0_(*t*) − *m*_1_(*t*)*}*, 0 ≤ *s* ≤ 1. Local approximations apply when *m*_0_(*t*) and *m*_1_(*t*) are close to a common mean 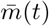.

With these likelihood-induced weights, the spectral separability in equation (5) is

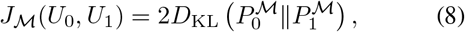

where 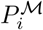 is the distribution of the downstream observation vector implied by *U*_i_ under observation model . This representation connects the spectral separability criterion to a likelihood-based testing bound.

For any decision rule *ϕ*, define the total testing error

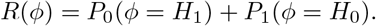

For two simple hypotheses,

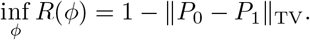

Here, ∥ · ∥_TV_ denotes the total variation distance. Combining equation (8) with Pinsker’s inequality gives

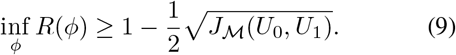

Consequently, for any target total testing error level *α*∈ (0, 1),

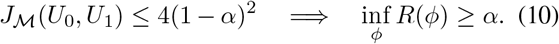

When this condition holds, no statistical procedure can reliably distinguish the two upstream trajectories from the downstream observations alone at the specified error criterion.

## Results

### Empirical delays impose strong low-pass filtering

Figure 1A–F illustrates how delayed observation limits the temporal resolution of epidemic surveillance in a data-informed example. We now ask whether this mechanism holds across published epidemiological delay distributions.

We evaluated the theoretical predictions using published epidemiological delay distributions spanning both biological delays (exposure-to-onset) and surveillance delays (symptom-onset-to-report) (Fig. 2A,D). These distributions cover diverse diseases, surveillance settings, geographic regions, and study periods, allowing us to test whether the predicted attenuation pattern is common across realistic epidemiological delay processes.

**Figure 2:**
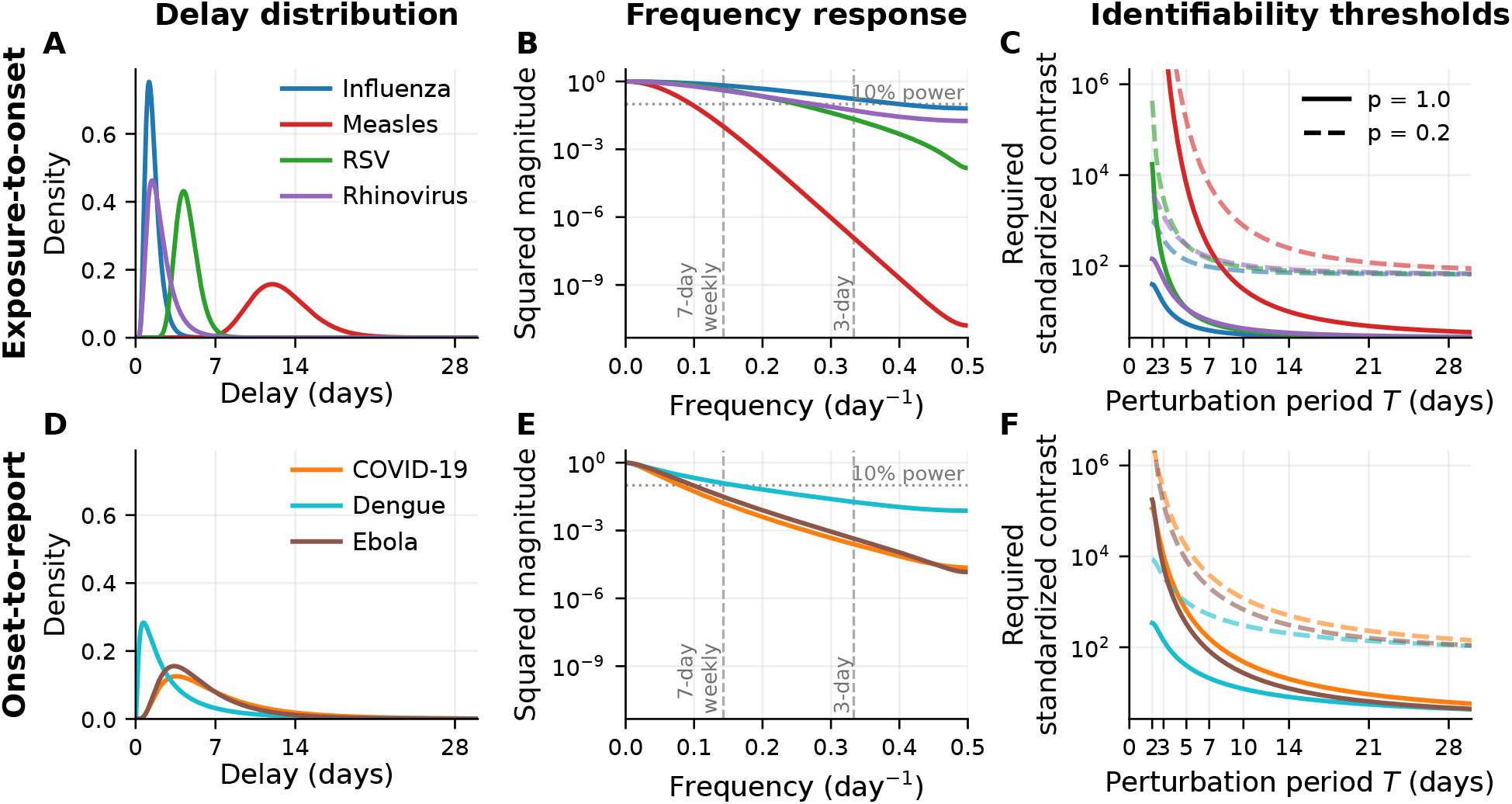
Frequency responses of representative epidemic delay distributions. (*A,D*) Published exposure-to-onset and onset-to-report delay distributions [26, 33, 3, 15]. (*B,E*) Corresponding squared magnitudes |*Ĝ*(*f* )| ^2^; dashed vertical lines mark 7-day and 3-day periods. (*C,F*) Minimum required standardized upstream contrast as a function of perturbation period *T* = 1*/f* under the single-frequency display. Solid and dashed curves correspond to conversion rates *p* = 1.0 and *p* = 0.2.

For each delay distribution *g*(*τ* ), we computed its frequency response *Ĝ* (*f* ) and squared magnitude |*Ĝ* (*f* )|^2^. Despite their heterogeneous epidemiological origins, all representative delay distributions exhibited rapidly decaying frequency responses (Fig. 2B,E), confirming that the examined epidemiological delays consistently behave as low-pass filters.

Frequencies corresponding to weekly and sub-weekly variation (i.e., *f* ≥ 1*/*7 cycles/day) fall in a strongly attenuated regime for many of the empirical delays. In some cases, especially for broader or more dispersed delay distributions, the power transmitted at weekly and faster frequencies is reduced by orders of magnitude relative to low-frequency components.

We next quantify how this attenuation translates into statistical detectability on a single-frequency standardized scale. Consider an upstream perturbation concentrated around a characteristic period *T*, corresponding to frequency *f*_0_ = 1*/T* . When the observation-model weight is diagonal in the Fourier basis, as for stationary Gaussian observation noise, the contribution of this perturbation to downstream separability is proportional to

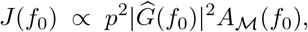

where *A*_*M*_(*f*_0_) denotes the upstream spectral contrast after standardization by the observation model. For stationary Gaussian observations, for example, 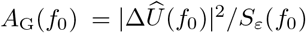. For count observations, the exact separability operator need not be diagonal in frequency; therefore Fig. 2C,F provide standardized single-frequency summaries of how delay filtering and observation variability affect contrasts at different temporal scales.

For a fixed target testing-error level *α*, the non-identifiability threshold derived in Methods implies that the minimum required standardized upstream contrast is

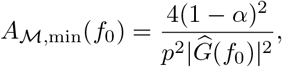

under this single-frequency representation.

Figure 2C,F visualize the minimum required standardized upstream contrast as a function of perturbation period *T* . Under the same observation window, shorter-period perturbations require larger standardized contrasts, reflecting the progressively stronger attenuation of high-frequency components by the delay kernel. Lower conversion rates further amplify this effect: reducing *p* from 1.0 to 0.2 increases the required contrast by a factor of 1*/p*^2^ = 25. Partial ascertainment therefore compounds the information loss caused by delay filtering.

Empirical epidemic delay distributions therefore impose a finite temporal resolution limit on upstream inference. The consequence is not only that delays smooth the observed data, but that they make short-timescale upstream variation statistically difficult to distinguish from observational variability. Perturbations occurring over several weeks may remain visible after delay convolution, whereas weekly or sub-weekly fluctuations often require unrealistically large standardized contrasts to be identifiable.

### Spectral identifiability constrains epidemiological inference

The consequence of limited identifiability is not ambiguity in trajectory reconstruction itself, but ambiguity in the scientific conclusions drawn from reconstructed trajectories. Whenever downstream data fail to identify part of the upstream process, reconstruction necessarily relies on information external to the data, including prior distributions, smoothness assumptions, regularization constraints, or mechanistic model structure [23, 31, 29]. These assumptions play an essential role in stabilizing ill-posed inverse problems. However, they also imply that weakly identified components of a reconstructed trajectory are determined jointly by the observed data and by the assumptions used to complete the inference problem.

We constructed frequency-constrained upstream trajectories from the same downstream COVID-19 incidence series. For each cutoff frequency *f*_c_, we estimated an upstream trajectory 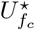 restricted to Fourier components satisfying *f* ≤ *f*_c_, while requiring its delay-convolved downstream projection to remain consistent with the same downstream observations (Methods; Appendix S2). We considered *f*_c_ = 1*/*60, 1*/*14, and 1*/*7 cycles/day, corresponding to perturbation periods of 60, 14, and 7 days, together with a no-cutoff reference trajectory. These trajectories illustrate how progressively admitting higher-frequency structure changes the range of admissible reconstructions.

To visualize this uncertainty, we computed frequency-constrained non-identifiable perturbation bands around each reference trajectory. These bands consist of perturbations whose downstream effect remains below the non-identifiability threshold implied by the separability criterion *J* (Appendix S2). The resulting bands widened as progressively higher-frequency components were admitted, illustrating that the set of upstream trajectories compatible with the same downstream observations expands when weaker frequency constraints are relaxed (Fig. 1A).

In practice, reconstructed incidence curves are rarely the final inferential target. Instead, they are commonly used to derive epidemiological quantities such as epidemic peak timing, peak magnitude, and short-term growth rates. To examine how spectral identifiability translates into epidemiological inference, we evaluated these three representative quantities across the admissible reconstruction family. For visualization, peak timing was expressed as the normalized location of the epidemic peak within the study period, and peak magnitude was normalized by the largest reconstructed peak across the admissible reconstruction family (Fig. 1H).

Although all admissible reconstructions generated nearly identical downstream trajectories (Fig. 1C), they did not support the same epidemiological conclusions. Epidemic peak timing and peak magnitude remained remarkably consistent across the reconstruction family (Fig. 1H), whereas inferred seven-day growth-rate trajectories differed substantially (Fig. 1G). The definition of the seven-day log growth rate is

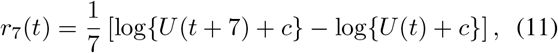

where *c >* 0 is a small numerical offset.

Unlike peak timing or peak magnitude, the seven-day growth rate is computed from local temporal differences. Temporal differencing emphasizes short-timescale variation (Appendix S3), causing growth-rate estimates to depend disproportionately on high-frequency components of the reconstructed trajectory. Under delayed observation, these are precisely the components that are most strongly attenuated by the delay kernel and least constrained by the downstream observations.

The differences in the inferred seven-day growth-rate trajectories (Fig. 1G) therefore reflect alternative completions of weakly identified high-frequency components rather than differences required by the downstream observations. Short-term growth-rate estimates are determined jointly by the information contained in the observations and by the assumptions used to reconstruct the weakly identified portion of the upstream trajectory.

Agreement in downstream goodness-of-fit does not imply equal support for all epidemiological conclusions. Reconstructions that are statistically indistinguishable at the observation level may nevertheless lead to different scientific inferences. The separability criterion distinguishes quantities constrained by the information contained in delayed observations from quantities that depend primarily on assumptions introduced to resolve weakly identified components of the inverse problem.

### Event impacts form reference-conditioned low-separability landscapes

Epidemiological events are often known independently of surveillance data. Mass gatherings are scheduled, temporary lockdowns are announced, and vaccination campaigns are documented administratively. The important inferential question is therefore often not whether an event occurred, but which transmission impacts are distinguishable from alternative event impacts after delay filtering and observational noise.

We characterized this uncertainty using reference-conditioned event identifiability landscapes. For a specified reference event profile, these landscapes map alternative combinations of event duration and transmission impact according to their downstream separability after epidemic propagation, delay convolution, and observation noise. We considered three stylized event families representing common epidemiological perturbations: a transient increase in transmission associated with a mass gathering, a transient reduction associated with a temporary lockdown, and a gradual reduction associated with a vaccination rollout [5, 17, 7]. The reference profiles corresponded to a 50% peak proportional change in *R*_t_, with characteristic durations of 3, 10, and 60 days, respectively.

We represented each event through a transmission multiplier *ρ*(*t*; *θ*) acting on a baseline reproduction number trajectory,

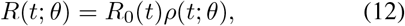

where *θ* = (*a, d*) indexes the event profile within each event family, *a* is the peak proportional change in *R*_*t*_, and *d* is the event duration or rollout timescale. The three transmission multipliers and their event-specific parameters are summarized in Table 2.

**Table 2:** Stylized event models used in the event-impact analysis. Each profile is indexed by *θ* = (*a, d*).

| Event type | Transmission multiplier | Reference profile |
| --- | --- | --- |
| Mass gathering | $1 + a h_d(t - t_0)$ | 50% increase, 3-day duration |
| Temporary lockdown | $1 - a h_d(t - t_0)$ | 50% reduction, 10-day duration |
| Vaccination rollout | $1 - a C_d(t)$ | 50% reduction, 60-day rollout |

The event window $h_d(t - t_0)$ is normalized to attain a maximum of one and is localized around the known event time $t_0$ . The rollout curve $C_d(t)$ increases monotonically from zero to one over a timescale controlled by $d$ .

To examine how alternative event profiles propagate through epidemic dynamics and delayed observation, we conducted a controlled synthetic experiment. We used controlled synthetic epidemics to isolate the effect of event profiles while holding all other epidemic and observation processes fixed. Across all event classes, we used the same baseline epidemic trajectory, generation interval distribution, renewal framework, delay kernel, conversion rate, and negative binomial observation model with a common dispersion parameter. Within each event family, every candidate profile was generated by perturbing the same baseline epidemic through the same renewal process, so that differences in *J*_event_ reflected only differences in the event profile under a common epidemic setting. Complete simulation details are provided in Appendix S4.

Incidence was generated through the renewal equation,

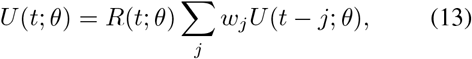

and mapped to the downstream mean through the same delay kernel used throughout the analysis, where *j* indexes discrete generation interval lags and *w*_j_ denotes the generation interval distribution.

For a specified reference event *θ*^∗^, we compared each candidate profile *θ* through the induced upstream contrast

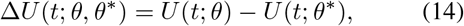

and the corresponding delayed downstream mean contrast

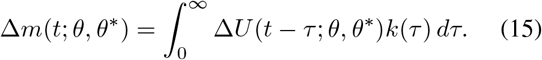

We then quantified event separability using the general likelihood weight defined in Methods,

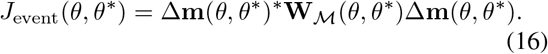

Here *M* denotes the downstream observation model, and **W**_*M*_(*θ, θ*^∗^) is the finite-contrast likelihood weight along the two candidate downstream mean trajectories. Equivalently, *J*_event_ can be written in the spectral operator form of Methods, with the delay transfer operator acting on 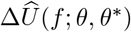. In the controlled examples shown below, we instantiate this general construction with a negative binomial observation model, because overdispersed count variation is common in epidemic surveillance data. The resulting landscape is necessarily reference-conditioned: changing *θ*^∗^ changes the position and geometry of its low-separability neighborhood, but not the procedure used to construct it.

The resulting landscapes exhibit extended low-separability neighborhoods surrounding each reference event (Fig. 3A,F,K). Within the cyan contour, the testing error bound derived in Methods guarantees that every statistical decision rule has total error at least *α* under the specified observation model. Candidate profiles in this region therefore cannot be reliably distinguished from the reference at the chosen error criterion.

**Figure 3:**
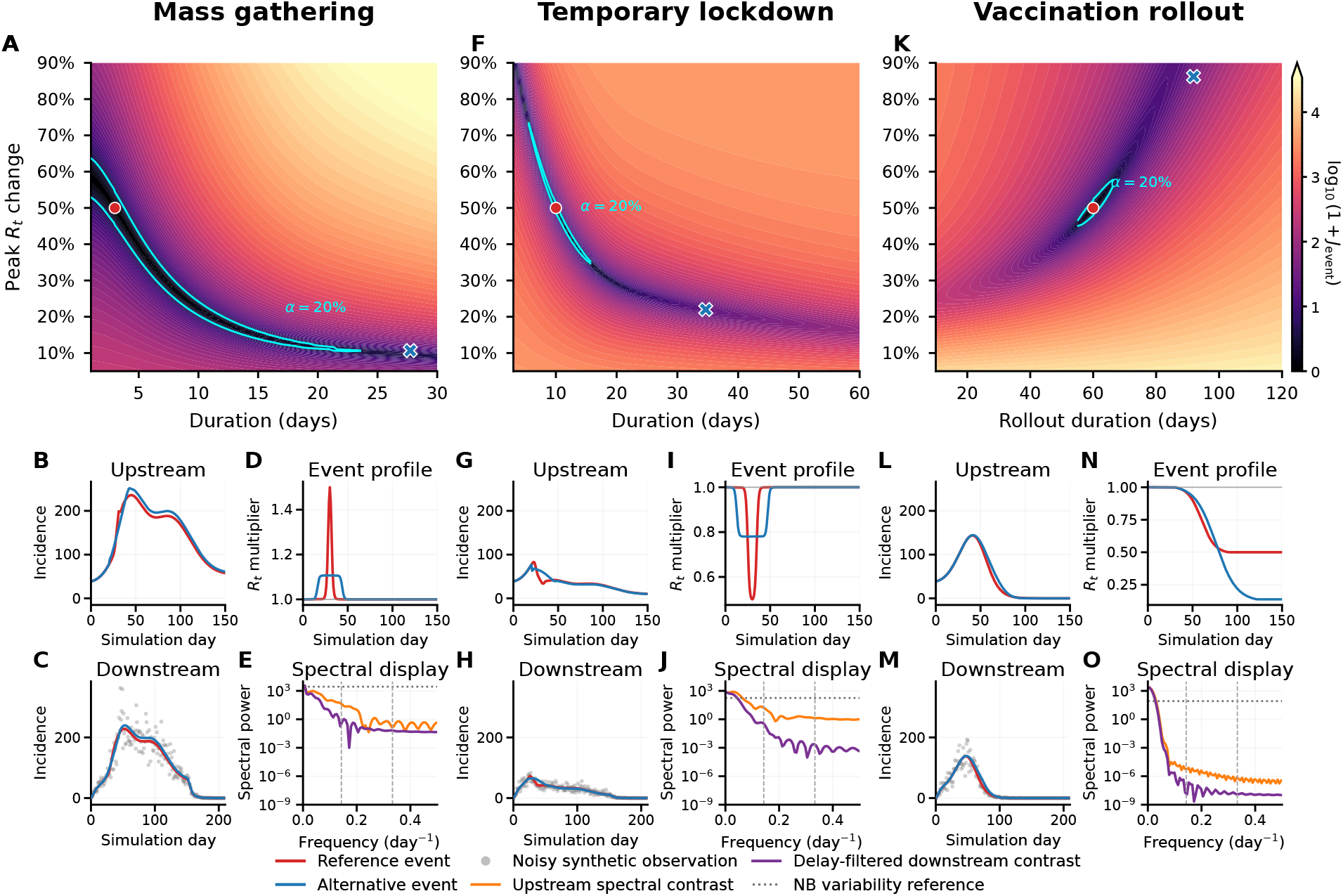
Reference-conditioned event-impact landscapes. Each column presents a reference-conditioned event-impact analysis for a mass gathering, temporary lockdown, or vaccination rollout. (*A,F,K*) Event identifiability landscapes over duration and peak proportional change in *R*_t_; color denotes log_10_(1 + *J*_event_), red circles mark reference profiles, and cyan contours mark *J*_event_ = 4(1 − *α*)^2^ for *α* = 0.20. (*B–E,G–J,L–O*) Reference and selected alternative profiles propagated through upstream incidence, transmission multipliers, downstream means, and spectra. Orange and purple curves show 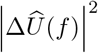 and 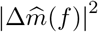, respectively; gray horizontal lines show one-sided negative binomial observation variability references. Displayed spectra omit the zero-frequency component.

The shape of each landscape reflects an event-specific tradeoff between transmission magnitude and event duration. For transient mass gatherings and temporary lock-downs, shorter but stronger perturbations can remain only weakly separated from longer but weaker perturbations. By contrast, vaccination rollout generates a more localized landscape because changes in rollout duration primarily alter lower-frequency epidemic dynamics.

To illustrate the practical implications of these landscapes, we selected one representative alternative for each event class outside the sufficient non-identifiability boundary while favoring large upstream trajectory contrast subject to remaining in a low-separability region (Appendix S4). The selected alternatives are representative members of the low-separability neighborhood. As shown in Fig. 3B–E,G–J,L–O, the reference and alternative profiles produce visibly different transmission multipliers and upstream incidence trajectories. After delay convolution, however, much of this contrast is attenuated, and the resulting downstream means remain similar relative to observational variability.

This loss of separability arose because much of the event-induced contrast was concentrated at temporal scales strongly reduced by the delay kernel. After delay filtering, the remaining downstream contrast was often comparable to typical negative binomial observation variability, even when the corresponding transmission histories and upstream incidence trajectories differed substantially. The spectral panels show this attenuation across temporal scales; *J*_event_ is evaluated from the likelihood-weighted downstream contrast defined in Methods.

Event occurrence and event-impact identifiability are therefore different inferential problems. Even when the timing and existence of an epidemiological event are known, delayed surveillance data may support a neighborhood of epidemiologically distinct transmission impacts rather than a unique estimate of event magnitude and duration. Reference-conditioned event identifiability landscapes quantify this inferential uncertainty under a specified epidemic and observation model.

## Discussion

Reporting delays are commonly understood as shifting and smoothing epidemic signals. Our results show that their more consequential effect is to impose a quantitative, frequency-dependent limit on what can be inferred from downstream surveillance data. By combining the transfer function of the delay process with conversion and observation-model-specific uncertainty, we characterize whether competing upstream epidemic trajectories can be statistically distinguished after they pass through the observation process. This perspective separates differences between epidemic histories that are supported by the transmitted information from those supplied mainly by reconstruction assumptions.

Across empirical delay distributions, distinguishability followed a consistent hierarchy across temporal scales and inferential targets. Slow epidemic variation is comparatively well preserved, whereas short-term variation, including weekly and sub-weekly fluctuations, can require substantially larger standardized upstream contrast to remain distinguishable after delayed observation. This loss of information does not affect all derived quantities equally. Features governed primarily by broad, low-frequency structure, such as the timing of a major epidemic wave, can remain comparatively stable across admissible reconstructions, while quantities based on local temporal differences, such as short-term growth rates, depend more strongly on weakly identified high-frequency components. The same principle extends to event attribution. Even when the occurrence and timing of a lockdown, mass gathering, or vaccination campaign are known, its transmission magnitude and temporal footprint may occupy an extended low-separability region rather than correspond to a uniquely identifiable parameterization. Identifiability is therefore not a binary property of an entire epidemic trajectory. It is specific to the temporal scale, epidemiological feature, and scientific comparison being considered.

These limits clarify the role of regularization, prior distributions, and mechanistic structure in epidemic reconstruction. Such assumptions are indispensable for stabilizing an ill-posed inverse problem and for producing epidemiologically coherent estimates. They cannot, however, recreate information that has been strongly suppressed by the observation process. When the likelihood provides little discrimination among alternative high-frequency trajectories, different priors, smoothing penalties, process models, or preprocessing choices may select different solutions while achieving nearly indistinguishable downstream fits. A reconstruction may consequently be numerically stable, have narrow conditional uncertainty, and closely reproduce the observed data without its fine-scale features being strongly identified by those data. Downstream goodness of fit, inferential precision, and statistical identification should therefore be treated as distinct properties.

The quantitative boundary between identifiable and weakly identifiable variation is conditional on the observation process through which epidemic information is transmitted. Among its components, uncertainty in the delay distribution is particularly consequential because the delay kernel directly determines the frequency-dependent attenuation of upstream variation. Our sensitivity analysis shows that moderate changes in the assumed median delay or dispersion can shift the identifiability boundary by orders of magnitude for rapid perturbations, whereas longer-timescale variation is generally less sensitive to the same uncertainty (Appendix S5; Fig. S1). Consequently, the temporal resolution limit should be evaluated across epidemiologically plausible delay specifications rather than under a single fixed kernel, particularly when inference concerns temporal scales near the resolution boundary.

Other components of the observation process—including ascertainment, observation noise, and the observation window—likewise influence the quantitative location of this boundary. Complementary observations, such as wastewater measurements, testing data, mobility or contact indicators, genomic information, and administrative records of interventions, can further reduce ambiguity by contributing information unavailable from a single delayed outcome stream. Their contribution should be interpreted as providing additional sources of identification, rather than as evidence that the original surveillance series itself contained the missing temporal information.

These results also bear on surveillance design and model evaluation. Shortening and narrowing reporting delays, increasing ascertainment, reducing measurement noise, and combining observations with complementary temporal responses do more than improve timeliness: they increase the range of epidemic variation that can be statistically resolved. Model evaluation should ask not only whether an estimate fits observed outcomes or produces a precise posterior, but also whether the particular epidemiological quantity being reported is identifiable under the assumed observation process and remains stable across plausible delay specifications and reconstruction assumptions. Making these limits explicit would shift epidemic inference away from presenting a single detailed latent history toward reporting which temporal scales, epidemiological features, and scientific conclusions are actually supported by the available surveillance system.

## Data Availability

All data produced are available online at: https://github.com/jingjtang/spectral-identifiability/tree/main

## Data Availability

The code used to generate all figures and analyses is publicly available at https://github.com/jingjtang/spectral-identifiability.

## Acknowledgments

We thank Valerie Ventura, Logan Brooks, Will Townes, and Ryan Tibshirani for helpful discussions.

This work was supported by the United States of America Department of Health and Human Services, Centers for Disease Control and Prevention, under award number U01IP001121 and contract number 75D30123C1590. Any opinions, findings, and conclusions or recommendations expressed in this material are those of the author(s) and do not necessarily reflect the views of the United States of America Department of Health and Human Services, Centers for Disease Control and Prevention.

## S1 Likelihood basis for the spectral separability criterion

This appendix derives the separability criterion used in Methods. We consider the simple hypotheses

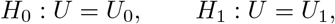

and let 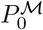 and 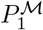 denote the corresponding laws of the downstream observations under observation model *M*.

### S1.1 Finite-window representation

Consider observations on a finite grid of *T* time points. Let **U**_i_ ∈ ℝ^T^ denote the sampled upstream trajectory under *H*_i_, and let

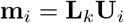

be the corresponding downstream conditional mean, where *k*(*τ* ) = *p g*(*τ* ). Here **L**_k_ is the finite-window linear delay-observation operator induced by the observation kernel *k*. In the main text, the Fourier representation is used to display the temporal scales affected by delay filtering. On finite windows, this representation should be understood under a convention in which boundary effects from the causal delay convolution do not drive the spectral quantities, such as analyzing a window with sufficient temporal margins or applying an explicit finite-window convention. Under the corresponding circular representation, this operator can be written as

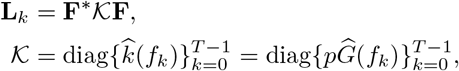

where **F** is the unitary discrete Fourier transform. Writing

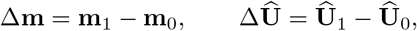

we have

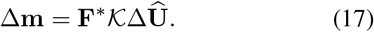

For a positive-semidefinite likelihood weight **W**_*M*_(*U*_0_, *U*_1_), the same criterion can be written in the downstream mean scale as

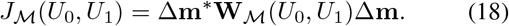

Combining equations (17) and (18) gives the spectral representation used in the main text,

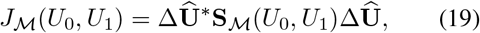

where

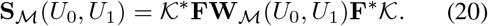

Thus the delay distribution enters through *K*, whereas the observation model enters through **W**_*M*_.

The likelihood weights derived below are directed: they correspond to the ordered comparison 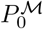 versus 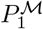. Reversing the hypotheses gives the analogous weights with *m*_0_ and *m*_1_ interchanged.

### S1.2 Testing error

For any decision rule *ϕ*, define the total testing error

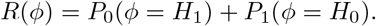

For two simple hypotheses,

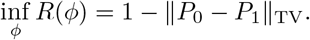

Pinsker’s inequality gives

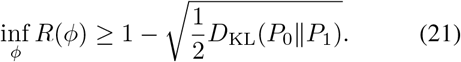

The following sections show that, for the observation models used in the paper,

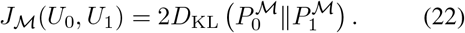

Substitution into equation (21) yields the common non-identifiability bound in the main text.

### S1.3 Gaussian observation model

Suppose that

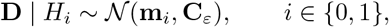

where **C**_ε_ is positive definite and common to the two hypotheses. The Gaussian laws differ only in their means, so

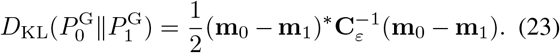

Set

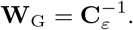

Then

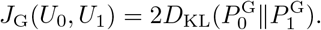

If the Gaussian noise is stationary, the Fourier basis diagonalizes the covariance:

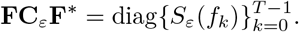

Consequently,

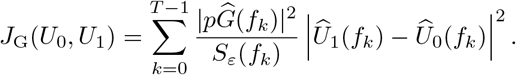

This is the exact frequency-wise expression for stationary Gaussian observation noise.

### S1.4 Likelihood curvature for count observations

For the count models, assume conditionally independent observations across time and strictly positive downstream means along the line segment between the two hypotheses:

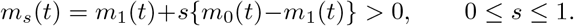

This condition excludes singular zero-mean cases; in numerical work it can be enforced by a small positive incidence floor.

Let *P*_m_ be a scalar observation law indexed by its mean *m >* 0, and suppose that the directed KL divergence can be written as the Bregman divergence generated by a twice continuously differentiable convex function *ψ*:

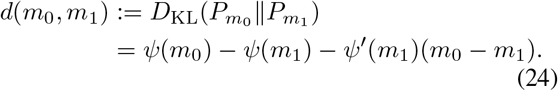

Taylor’s theorem with integral remainder gives

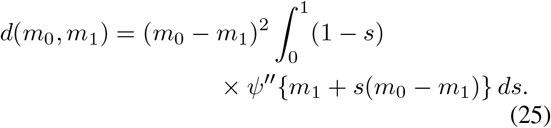

For a regular one-parameter exponential-dispersion family parameterized by its mean,

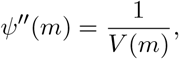

where *V* (*m*) = Var(*D* | *m*) is the variance function. Therefore

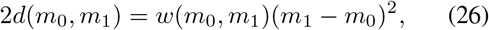

with exact finite-contrast likelihood weight

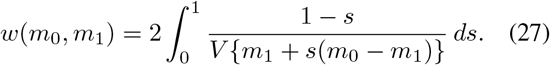

If *m*_0_ and *m*_1_ are close to a common reference value 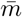, then

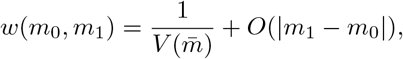

which recovers the inverse-variance Fisher-information weight as a local approximation.

### S1.5 Poisson observation model

Suppose that

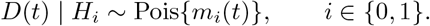

For scalar Poisson observations,

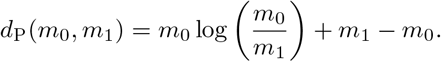

This is the Bregman divergence generated by

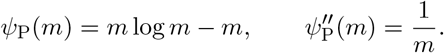

Thus *V*_P_(*m*) = *m*, and equation (27) gives

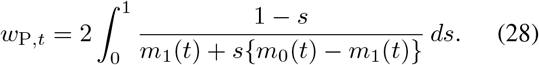

Let

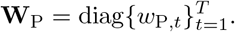

Conditional independence and equation (26) imply

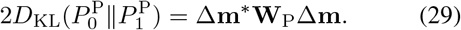

Therefore,

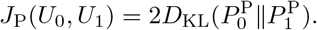

When the two mean trajectories are locally close,

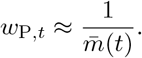

### S1.6 Negative-binomial observation model

Suppose that

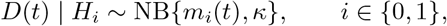

where the dispersion parameter *κ >* 0 is shared by the two hypotheses and

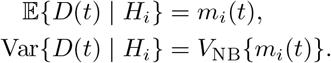

with

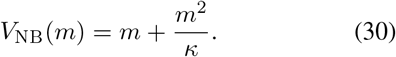

For fixed *κ*, the scalar directed KL divergence is the Bregman divergence generated, up to affine terms, by

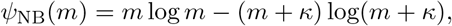

whose curvature is

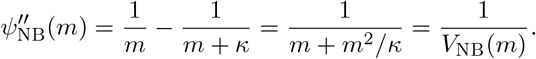

Equation (27) therefore gives

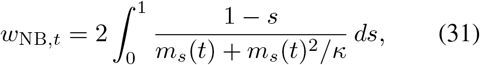

where *m*_s_(*t*) = *m*_1_(*t*) + *s{m*_0_(*t*) − *m*_1_(*t*)*}*. Let

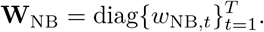

Conditional independence gives

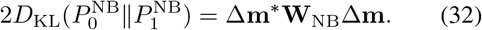

Therefore,

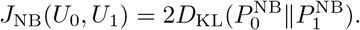

When the two mean trajectories are locally close,

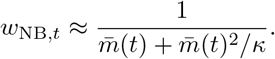

### S1.7 Common non-identifiability threshold

For each observation model above,

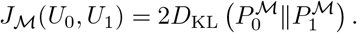

Substituting this identity into equation (21) gives

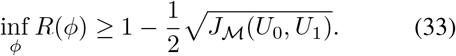

Thus, for any target total testing error *α* ∈ (0, 1),

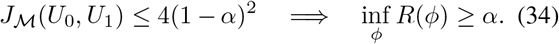

The threshold is common across observation models because *J*_*M*_ is defined using the likelihood geometry. The observation model affects distinguishability through **W**_*M*_, and the delay process affects distinguishability through *K*.

## S2 Construction of the combined illustrative example in Figure 1

Figure 1 provides a data-informed illustration of how delay filtering limits the temporal resolution of epidemic surveillance and how this limitation propagates to epidemiological features. The figure is intended to illustrate the observation and separability framework rather than to provide a substantive reconstruction of the U.S. epidemic.

### S2.1 Observed downstream incidence

Daily U.S. confirmed-case incidence was obtained from the Johns Hopkins University Center for Systems Science and Engineering COVID-19 data stream [12], accessed through the Delphi Epidata API [16]. We used the national-level `jhu-csse/confirmed_incidence_num` signal.

The complete series from April 1, 2020, through June 30, 2021, was used to estimate the smoothed downstream mean, construct the candidate upstream trajectories, and perform convolution, thereby preserving the pre-window history required for observations near the beginning of the displayed interval. The spectral quantities, non-identifiable perturbation bands, and epidemiological feature summaries in Fig. 1 were evaluated over the displayed period from September 1, 2020, through March 31, 2021.

### S2.2 Smoothed downstream mean and observation model

To separate broad epidemic variation from day-to-day observational fluctuation, we estimated a smooth downstream mean 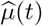 using second-order trend filtering. The effective degrees of freedom were selected over a pre-specified grid. Conditional on each candidate smooth mean, Gaussian, Poisson, and negative-binomial observation models were fitted, and the combination of trendfilter complexity and observation model was selected by the Akaike information criterion (AIC). The selected observation model was negative binomial for the displayed U.S. case series. Its fitted variance function was used to construct the local likelihood weights for the perturbation bands and the observation-variability reference shown in Fig. 1F.

### S2.3 Delay distribution

The delay kernel was based on the published COVID-19 symptom-onset-to-case-report distribution reported by Zhang et al. [33]. A lognormal representation was converted to a discrete daily probability mass function by integrating the fitted distribution over consecutive one-day intervals. The kernel was truncated at 60 days and renormalized to sum to one. The confirmed-case example used conversion rate *p* = 1.

### S2.4 Frequency-constrained reference trajectories

For each cutoff frequency *f*_c_, we represented the upstream trajectory in a real Fourier basis 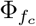 containing the constant component and sine and cosine components with frequencies satisfying 0 ≤ *f* ≤ *f*_c_. The cutoff-specific reference trajectory 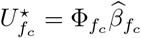 was obtained by solving

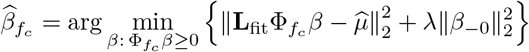

where **L**_fit_ is the finite-window linear delay-observation matrix mapping upstream incidence to the downstream mean over the fitting window, 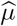 is the fitted smooth downstream mean, *β*_−0_ denotes all nonconstant Fourier coefficients, and *λ* = 10^−6^ is a small ridge penalty used for numerical stability. Figure 1A,C,G,H show cutoff frequencies *f*_c_ = 1*/*60, 1*/*14, and 1*/*7 cycles/day, corresponding to characteristic periods of 60, 14, and 7 days. We also constructed a no-cutoff reference by setting *f*_c_ = 0.5 cycles/day, the Nyquist frequency for daily data when present.

### S2.5 Local non-identifiable perturbation bands

The shaded bands in Fig. 1A use the local likelihood approximation described in Appendix S1. For each cut-off *f*_c_, perturbations were restricted to the same Fourier subspace as the corresponding reference trajectory. Let 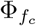 denote this basis on the displayed window, **L**_disp_ the displayed-window linear delay-observation matrix, and **W**_loc_ the diagonal matrix of local likelihood weights under the selected observation model. The local downstream separability of a perturbation 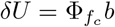 is

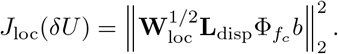

Equivalently, writing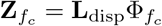,

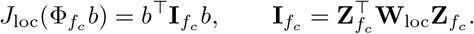

For *α* = 0.20, we used the non-identifiability threshold *J*_α_ = 4(1 − *α*)^2^ = 2.56, giving the perturbation ellipsoid

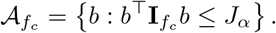

At each displayed time point *t*, the pointwise perturbation radius was computed as

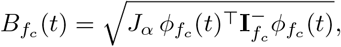

where 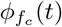 is the vector of Fourier-basis values at time *t*, and 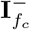 is the Moore–Penrose inverse used in the numerical calculation, equal to the ordinary inverse when 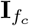 is full rank. The displayed band is

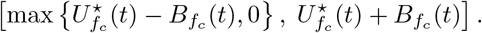

This band is a pointwise envelope of the local perturbation ellipsoid. The perturbation attaining the endpoint may differ across time points.

### S2.6 Spectral display and observation-variability reference

The spectral quantities in Fig. 1D–F were computed over the displayed period using the same one-sided discrete Fourier transform and frequency grid. Tilde notation denotes the corresponding discrete Fourier coefficient on this displayed window; for example, 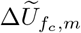 is the coefficient of 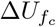 at frequency *f*_m_. These panels display how upstream spectral contrast is attenuated before it appears in the downstream mean; separability itself is evaluated using the likelihood-weighted quadratic form described in Methods. Panel D shows, for each cutoff-specific reference trajectory, the upstream spectral contrast relative to the no-cutoff reference,

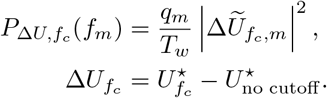

where *T*_w_ is the displayed-window length and *q*_m_ is the standard one-sided multiplicity weight. In the displayed spectra, the zero-frequency component is omitted because it represents mean-level contrast rather than temporal variation. For the one-sided display, *q*_m_ = 2 for positive interior frequencies and *q*_m_ = 1 for boundary frequencies, including the zero-frequency component and the Nyquist frequency *f* = 0.5 cycles/day when it is present. Panel E shows the squared delay attenuation 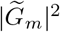, where 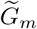 is the discrete Fourier coefficient of the delay kernel at frequency *f*_m_. Panel F shows the corresponding downstream-mean spectral contrast computed from the displayed portions of the full delay-convolved reference means,

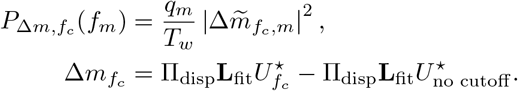

Here Π_disp_ denotes restriction to the displayed period, so the downstream spectra are computed after full-window convolution and then restricted to the displayed period.

The horizontal line in Fig. 1F is a visual reference for the scale of observation variability on the one-sided frequency scale. For Poisson and negative-binomial observations, we compute local likelihood weights *w*_t_ at the fitted downstream mean and display

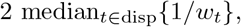

where the median is taken over the displayed period. The factor of two accounts for the pairing of positive- and negative-frequency coefficients at interior frequencies in the one-sided display. Separability calculations use the likelihood-weighted quadratic form described in Methods.

## S3 Epidemiological feature summaries for the combined illustrative example

Here we describe the epidemiological feature summaries in Fig. 1G,H and explain why the seven-day growth-rate summary is especially sensitive to weakly identified high-frequency components.

### S3.1 Epidemiological feature summaries

We evaluated three quantities derived from each displayed upstream reference trajectory: epidemic peak timing, epidemic peak magnitude, and the seven-day log growth rate. All feature summaries were calculated over the displayed interval from September 1, 2020, through March 31, 2021.

### S3.1.1 Peak timing and peak magnitude

For each displayed upstream reference trajectory, we first formed a reflected 14-day moving average,

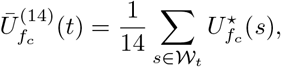

where *W*_t_ is the 14-day smoothing window used for the displayed trajectory, with reflection at the boundaries of the finite interval. The peak index was defined as

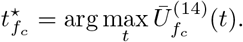

For visualization, relative peak timing was calculated as

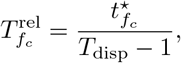

where *T*_disp_ is the number of days in the displayed analysis interval and 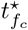 is indexed from zero within that interval.

The peak magnitude was evaluated from the unsmoothed cutoff-specific reconstruction at the peak index selected from the smoothed trajectory,

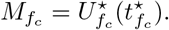

Relative peak magnitude was then defined as

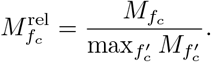

The common 14-day smoothing used to locate the epidemic peak reduces the sensitivity of the peak summaries to isolated daily fluctuations while preserving the broad epidemic-wave structure.

### S3.1.2 Seven-day log growth rate

For each displayed upstream reference trajectory, we calculated the seven-day log growth rate

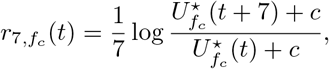

where *c* = 1 is a numerical offset used to avoid taking the logarithm of zero. Values within the final seven days of the displayed interval do not have a forward seven-day comparison and were filled from the nearest available growth-rate values before applying the common smoothing step. The resulting growth-rate trajectory was smoothed using a reflected five-day moving average. This final smoothing was applied identically to all displayed reference trajectories.

### S3.2 Spectral sensitivity of the seven-day growth rate

The seven-day growth rate is nonlinear in *U* (*t*), but its local spectral sensitivity can be seen through a first-order perturbation. Let

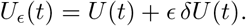

Differentiating *r*_7_(*t*) with respect to *ϵ* at *ϵ* = 0 gives

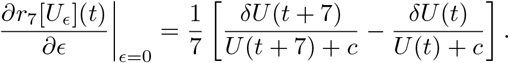

If the denominator varies slowly over a local interval, so that

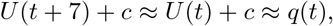

then

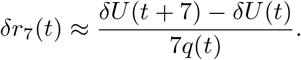

The corresponding temporal-difference operator has frequency response

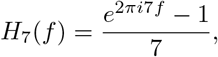

with magnitude

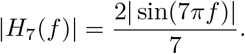

For low frequencies,

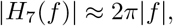

so the operator suppresses nearly constant components and increases its sensitivity as frequency moves away from zero. Relative to summaries of the overall level or broad epidemic-wave shape, the growth-rate calculation therefore places greater emphasis on short-timescale differences in the reconstructed trajectory.

The five-day moving average subsequently applied to the estimated growth rate attenuates the highest-frequency numerical variation, but it does not remove the underlying dependence of local growth estimates on upstream components that are more weakly transmitted through the delay process.

Peak timing and peak magnitude are nonlinear global functionals and do not possess a fixed linear transfer function analogous to *H*_7_(*f* ). Their relative stability in Fig. 1H is therefore an empirical property of the admissible reconstruction family considered here, rather than a universal theorem that peak summaries are always identifiable. The analysis shows that, for this epidemic series and these reconstruction classes, the broad peak structure is considerably more stable than the local growth-rate trajectory.

## S4 Controlled synthetic event-generation framework

This section describes the controlled synthetic framework used to construct the event-identifiability landscapes in Fig. 3. The objective of these simulations was to isolate how different event parameterizations propagate through epidemic dynamics, delay filtering, and observational noise under a common epidemic setting.

### S4.1 Baseline epidemic

All event classes were evaluated using the same baseline epidemic trajectory. The baseline incidence curve was defined as a smooth bimodal epidemic,

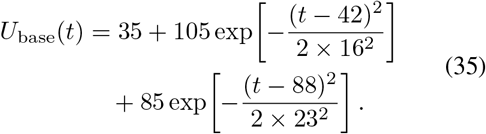

This baseline was evaluated over a simulation window consisting of 30 pre-event days and 120 post-event days. The transient-event profiles were centered at day 30; the vaccination-rollout profile began at day 30.

The corresponding baseline reproduction-number trajectory was inferred from the renewal equation,

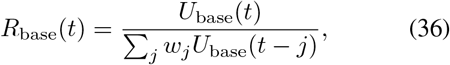

where *w*_*j*_ denotes the discrete generation-interval distribution. This baseline *R*_*t*_ trajectory served only as the reference transmission process on which event perturbations were imposed.

### S4.2 Generation interval

Transmission dynamics followed the discrete renewal model described in the main text. The generation interval was represented by a Gamma distribution with mean 5 days and standard deviation 2 days, discretized over lags of 1–21 days and normalized to sum to one.

### S4.3 Event parameterization

Each event profile modified the baseline reproduction number through

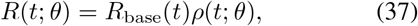

where *θ* = (*a, d*) indexes the event profile within each event family, *a* is the peak proportional change in *R*_t_, and *d* is the event duration or rollout timescale.

Three stylized transmission perturbations were considered.

#### Mass gathering

A transient increase in transmission was represented as

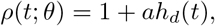

where *h*_d_ is a smoothed finite-duration window normalized to have unit maximum.

#### Temporary lockdown

A transient reduction in transmission was represented as

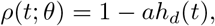

using the same normalized event window.

#### Vaccination rollout

Vaccination was represented by a monotone rollout curve,

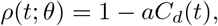

where *C*_*d*_(*t*) is a normalized logistic coverage curve increasing from 0 to 1 over the rollout period.

The event window *h*_*d*_ was constructed using two opposing logistic transitions,

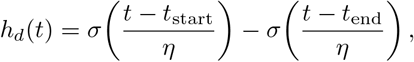

where *t*_start_ = *t*_0_ − *d/*2, *t*_end_ = *t*_0_ + *d/*2, and *η* = 1 day is the transition width used in the displayed simulations. The logistic function is

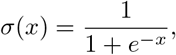

and *h*_d_ was then normalized to have unit maximum.

The vaccination rollout curve was generated using a logistic function centered at the midpoint of the rollout period and linearly rescaled so that

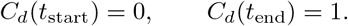

### S4.4 Renewal simulation

For every candidate event profile,

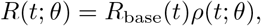

the corresponding incidence trajectory was generated recursively using

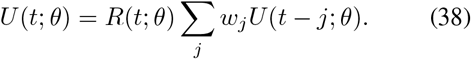

The first generation-interval window was initialized using the baseline trajectory. Consequently, within each event family every candidate profile was generated by perturbing the same baseline epidemic through the same renewal process. Differences across the event-identifiability landscape therefore arose solely from differences in the event parameterization under a common epidemic and observation setting.

### S4.5 Observation model

The downstream conditional mean was obtained through convolution with a common reporting-delay distribution,

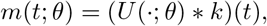

where *k*(*τ* ) = *p g*(*τ* ) and *p* = 1 in all controlled simulations.

The delay distribution was the same published COVID-19 symptom-onset-to-case-report kernel used in Fig. 1, based on [33]. The fitted lognormal distribution was discretized into daily probabilities, truncated at 60 days, and renormalized to sum to one. Thus the event profiles and epidemic trajectories were synthetic controlled examples, whereas the reporting-delay component was anchored to the empirical surveillance delay used in the data-informed illustration.

Full linear convolution was retained when constructing downstream means,

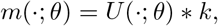

and the statistical comparison was evaluated on the observed upstream time window. For spectral visualization, we computed one-sided spectra for the upstream contrast Δ*U* and for the delay-filtered downstream mean contrast Δ*m* on this same finite window. The zero-frequency component was omitted from the displayed spectra because it represents mean-level contrast rather than temporal variation. This finite-window calculation avoids relying on a circular-convolution approximation for the event figure.

### S4.6 Negative-binomial observations

Illustrative observations were generated independently from

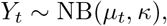

where

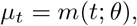

and

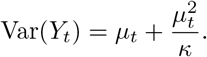

A common dispersion parameter

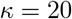

was used throughout all simulations.

One synthetic realization was generated from each reference event and displayed in Fig. 3 to illustrate the scale of observational variability. All identifiability calculations were performed using the corresponding downstream conditional means rather than the sampled observations.

### S4.7 Reference event profiles

The reference event profiles corresponded to a peak proportional 50% change in *R*_t_.

The reference durations were

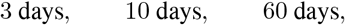

for the mass gathering, temporary lockdown, and vaccination rollout, respectively.

Candidate event landscapes were evaluated over

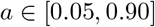

and event-specific duration grids covering

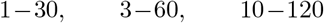

days for the three event classes.

### S4.8 Construction of the event-identifiability landscapes

For every point on the parameter grid, the corresponding incidence trajectory was generated through the renewal model and mapped to the downstream conditional mean. Each point was then compared with the specified reference profile for that event family. Let *θ*^∗^ denote the reference profile and *θ* denote a candidate profile. To match the notation in Methods, define *U*_0_(*t*) = *U* (*t*; *θ*^∗^), *U*_1_(*t*) = *U* (*t*; *θ*), and *m*_i_ = (*U* _i_. ∗ *k*), *i*∈ { 0, 1 }.

The event-identifiability landscape was then computed using the general finite-contrast form

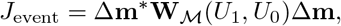

where

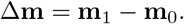

Thus every point in the landscape represents the downstream separability between a candidate event profile and the specified reference profile. In Fig. 3, was instantiated as the negative-binomial observation model described above. The weight matrix **W**_*M*_(*U*_1_, *U*_0_) was diagonal in time, with entries given by the exact finite-contrast negative-binomial likelihood weights derived in Appendix S1; this ordering corresponds to 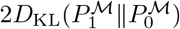. For the spectrum panels, we plotted the upstream spectral contrast | *Δ*Û (*f* )|^2^ and the corresponding delay-filtered downstream mean contrast 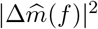, where Δ*U* = *U*_1_ − *U*_0_. The horizontal reference line was set to 2 median_t_ *m*_0_(*t*) + *m*_0_(*t*)^2^*/κ*, where *m*_0_(*t*) is the reference downstream mean on the observed window. This line gives a one-sided visual scale for typical negative-binomial observation variability.

### S4.9 Selection of illustrative alternatives

For each event class, one representative alternative was selected automatically for visualization.

Candidates were restricted to those lying outside the sufficient non-identifiability region,

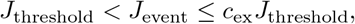

where *J*_threshold_ = 4(1 − *α*)^2^ with *α* = 0.20, while satisfying minimum parameter separation from the reference and excluding points near the parameter-space boundary. We used *c*_ex_ = 3, 8, and 18 for the mass-gathering, temporary-lockdown, and vaccination-rollout examples, respectively. The larger values for the latter two examples make the displayed reference–alternative contrasts visually interpretable; the event-identifiability landscapes themselves were computed from the same *J*_event_ criterion for all candidate profiles.

Among the remaining candidates, the displayed example was chosen to maximize the relative upstream root-mean-square difference from the reference trajectory. This procedure produces examples with visible differences in transmission dynamics while keeping the alternatives in a low-separability neighborhood outside the conservative non-identifiability boundary.

### S5 Sensitivity Analysis

The preceding analyses treat the delay distribution *g*(*τ* ) as known. In practice, epidemiological delay distributions are estimated from finite data and may be affected by sampling variability, right truncation, and reporting incompleteness. Uncertainty in the location or dispersion of the delay distribution changes its frequency response |*Ĝ* (*f* ) | ^2^, and therefore changes the amount of upstream spectral variation that remains distinguishable after delayed observation.

We examined how the single-frequency identifiability boundary varies across plausible delay distributions and under local misspecification of empirically estimated delay parameters. For an upstream perturbation with period *T*, the corresponding frequency is

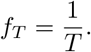

This appendix focuses on the delay-dependent part of the single-frequency threshold. On a standardized contrast scale, the non-identifiability threshold is

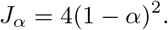

The minimum required standardized upstream contrast for statistical distinguishability is therefore

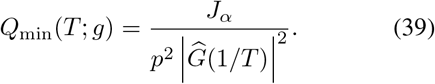

We set *α* = 0.20, giving *J*_α_ = 2.56, and fixed the conversion rate at *p* = 1. Larger values of *Q*_min_ indicate that greater likelihood-standardized upstream contrast is required for distinguishability.

We evaluated Eq. (39) for perturbation periods of 3, 7, and 14 days across a family of lognormal delay distributions. Each delay distribution was parameterized by its median delay and dispersion factor,

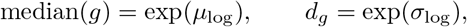

where *µ*_log_ and *σ*_log_ are the mean and standard deviation of the logarithm of the delay, consistent with the parameterization used by Lessler et al. [26]. The resulting surfaces in Fig. S1A–C show

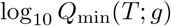

over the delay-parameter space.

The required standardized contrast is generally larger for broader delay distributions and for shorter-period perturbations. This pattern follows directly from the stronger attenuation of high-frequency variation by dispersed delay kernels. The 3-day perturbation is therefore substantially more sensitive to the assumed delay distribution than the 7-day and 14-day perturbations. In some regions of parameter space, relatively modest changes in the delay parameters produce large multiplicative changes in the contrast required for distinguishability.

To place these theoretical surfaces in an epidemiological context, we overlaid representative empirical delay estimates. These included exposure-to-symptom-onset distributions and symptom-onset-to-report distributions from several infectious diseases. The empirical estimates occupy different regions of the delay-parameter space and consequently imply different levels of attenuation at the same perturbation frequency. Long and dispersed reporting delays tend to require substantially greater upstream contrast for distinguishing rapid variation than shorter and more concentrated delays.

We next quantified local sensitivity around each empirical delay estimate. Let *M*_0_ and *d*_g,0_ denote the original median delay and dispersion factor, respectively. For median misspecification, we evaluated

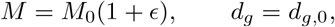

whereas for dispersion misspecification we evaluated

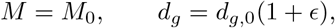

with

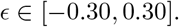

Parameter combinations yielding an invalid lognormal dispersion factor, *d*_g_ ≤ 1, were excluded.

For each perturbed delay distribution, we recomputed the theoretical single-frequency threshold in Eq. (39). Sensitivity was summarized relative to the threshold under the original empirical delay estimate:

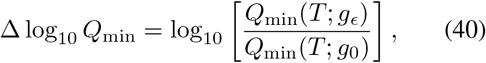

where *g*_0_ is the original delay distribution and *g*_ϵ_ is the distribution obtained after perturbing either its median or dispersion. Thus,

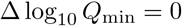

corresponds to the original empirical estimate, positive values indicate a higher required standardized contrast, and negative values indicate a lower required standardized contrast.

Fig. S1D–F shows the effects of median-delay misspecification. The direction and magnitude of the response depend on both the empirical delay distribution and the perturbation period. The largest changes generally occur for 3-day variation, whereas the required contrast for 14-day variation is comparatively less sensitive to moderate changes in the assumed median delay.

Fig. S1G–I shows the corresponding effects of dispersion misspecification. Increasing the dispersion factor generally increases the required standardized contrast because a broader delay distribution more strongly attenuates rapid upstream variation. This effect is again most pronounced for the 3-day perturbation and progressively weaker for the 7-day and 14-day perturbations.

Overall, uncertainty in the delay distribution changes the quantitative location of the single-frequency identifiability boundary but does not alter the principal qualitative conclusion: rapid upstream variation is substantially more difficult to distinguish than slower variation after delayed observation. Inference concerning short-timescale epidemic dynamics is therefore especially sensitive to uncertainty in the assumed delay kernel and should be interpreted together with uncertainty in the estimated delay parameters.

**Figure S1:**
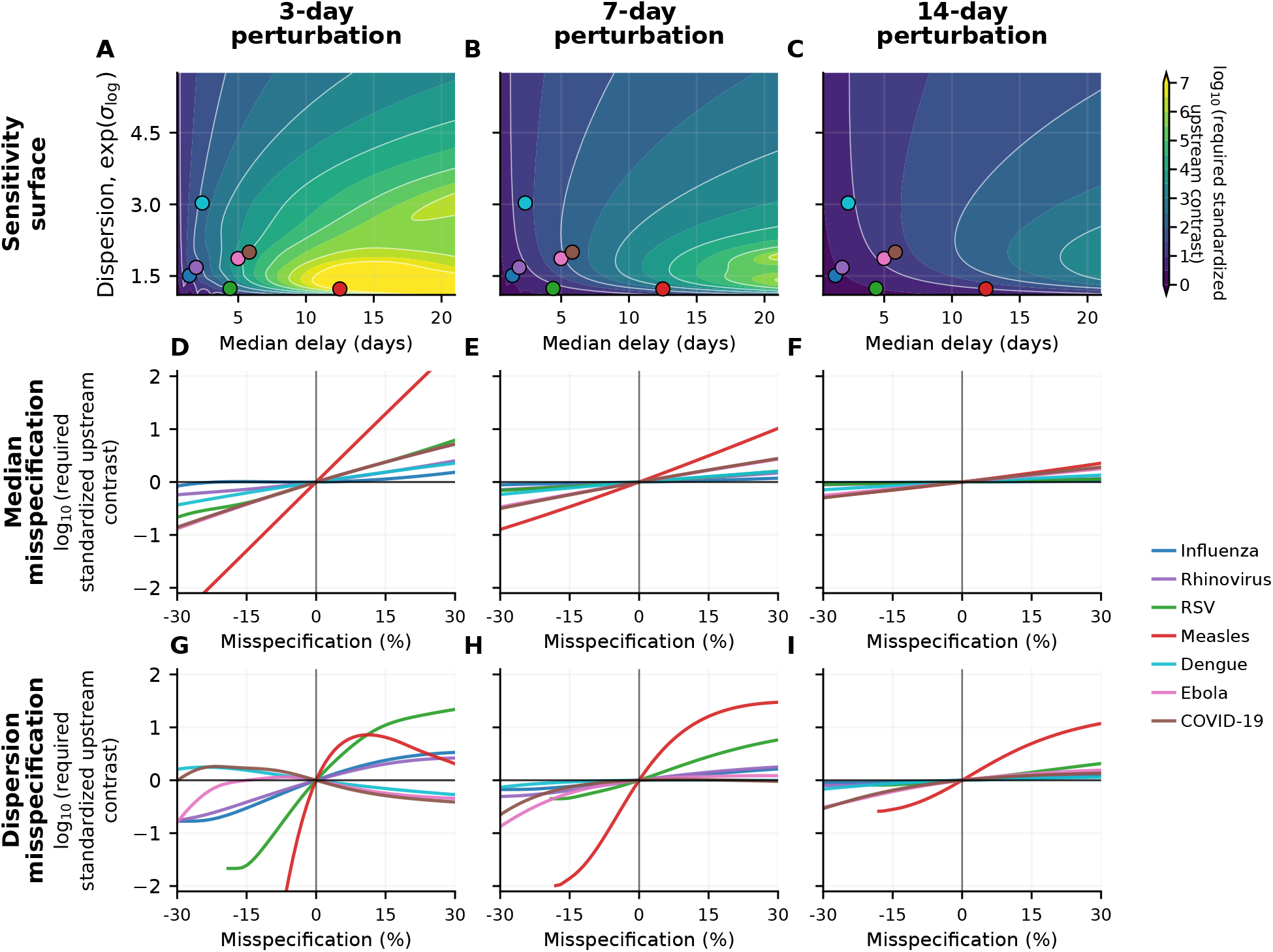
Sensitivity of single-frequency identifiability to delay-distribution parameters. (*A–C*) log_10_ *Q*_min_ across lognormal delay distributions parameterized by median delay and dispersion factor, shown for perturbation periods *T* = 3, 7, and 14 days with *α* = 0.20 and *p* = 1. Colored points denote representative empirical exposure-to-onset and onset-to-report delay estimates. (*D–F*) log_10_(*Q*_min_*/Q*_min,0_) after perturbing the median delay by up to *±* 30% while holding dispersion fixed. (*G–I*) Corresponding relative change after perturbing the dispersion factor while holding the median delay fixed.

